# Atherosclerosis, Intracranial Aneurysms, and Intermediate Biomarkers: Real-World Observational and Mendelian Randomization Research

**DOI:** 10.1101/2025.04.11.25325705

**Authors:** Wei Liu, Zhaoxu Zheng, Chenglong Liu, Yuanren Zhai, Shuang Wang, Liangran Huang, Rong Wang, Yan Zhang, Peicong Ge, Dong Zhang

**Affiliations:** Department of Neurosurgery, Beijing Tiantan Hospital, Capital Medical University, Beijing, China; China National Clinical Research Center for Neurological Diseases, Beijing, China, Center of Stroke, Beijing Institute for Brain Disorders, Beijing, China; Department of Neurosurgery, Beijing Hospital, National Center of Gerontology, Beijing, China; Institute of Geriatric Medicine, Chinese Academy of Medical Sciences, Beijing, China

**Keywords:** Atherosclerosis, Intracranial Aneurysms, Mendelian Randomization, risk factors, matrix metalloproteinase, Stenosis

## Abstract

**Background:** The causal relationship between atherosclerosis and the development of aneurysms remains controversial. This study aims to analyze the genetic predictive association between atherosclerosis and intracranial aneurysms, as well as to explore intermediate biomarkers that may mediate the causal relationship between these two conditions.

**Methods:** This study utilized head imaging data for a cross-sectional analysis, investigating the prevalence of cerebral atherosclerosis and intracranial aneurysms in 13,739 patients. We performed Mendelian randomization analysis on two samples to explore the genetic predictive association between atherosclerosis and intracranial aneurysms. Furthermore, we examined the role of the matrix metalloproteinase family as an intermediary biomarker in the causal relationship between these two conditions and validated biomarker in serum and in situ samples from clinical patients.

**Results:** Cross-sectional study revealed that patients with intracranial aneurysms exhibited a higher prevalence of cerebral atherosclerosis compared to controls (14.155% vs. 7.069%, P<0.001). Two-sample Mendelian randomization analysis demonstrated that genetically predicted peripheral atherosclerosis significantly increased the risk of intracranial aneurysms, encompassing both unruptured (OR_IVW_ = 1.711, P_IVW_ = 0.002) and ruptured intracranial aneurysms (OR_IVW_ = 1.533, P_IVW_ = 9.41×10^-4). Mendelian randomization further indicated that peripheral atherosclerosis leads to elevated circulating MMP12 levels (β_IVW_ = 0.083, P_IVW_ = 0.008), which were subsequently associated with increased risks of unruptured (OR_IVW_ = 1.136, P_IVW_ = 0.010) and ruptured intracranial aneurysms (OR_simple-mode_ = 1.217, P_simple-mode_ = 0.038). ELISA assays confirmed elevated MMP12 levels in the plasma of patients with carotid artery stenosis and intracranial aneurysms. In situ analyses revealed upregulated MMP12 expression in both plaques and intracranial aneurysm tissues.

**Conclusion:** Our research clarifies the causal relationship between atherosclerosis and intracranial aneurysms, identifies and validates MMP12 as a key biomarker, and provides new methods and evidence to explain the association between these two diseases.

**What is already known on this topic:** Previous studies have suggested a significant association between atherosclerosis and the development of aneurysms.

**What this study adds:** This study provides robust evidence and causal inferences regarding this relationship, identifying intermediate biomarkers of the diseases.

**How this study might affect research, practice or policy:** These findings suggest that preventing atherosclerosis and controlling MMP12 levels may help reduce the risk of intracranial aneurysms.

## Background

Intracranial aneurysm, a common cerebrovascular disease, typically manifests as an enlargement or protrusion of cerebral blood vessels. Research indicates a prevalence of 1%-2% in the general population^1^, with a cross-sectional study in Shanghai revealing a 7% detection rate in adults aged 35 to 75 years through comprehensive brain magnetic resonance angiography (MRA) screening^2^. Aneurysm rupture, which often leads to nontraumatic subarachnoid hemorrhage with high mortality rates, is primarily attributed to inflammation, matrix degradation, thrombosis, and hemodynamic factors according to animal model studies^3,4^. Risk factors for intracranial aneurysms include smoking, hypertension, substance abuse, hereditary connective tissue disorders, histopathies, and polycystic kidney disease^5^.

Both atherosclerosis and aneurysms are complex diseases characterized by inflammatory cell infiltration, matrix degradation, and arterial wall thrombosis. A case-control study found that patients with risk factors for atherosclerosis are more likely to develop middle cerebral artery aneurysms^6^. Furthermore, a retrospective cross-sectional study observed that unruptured intracranial aneurysm patients with atherosclerotic plaques in the middle cerebral artery exhibited more pronounced aneurysm wall enhancement, suggesting a potential association between the presence of plaques and intracranial aneurysm characteristics^7^. A study using fluid-structure interaction analysis to investigate the interaction between blood and vascular tissue caused by atherosclerotic occlusion found that hemodynamic changes induced by atherosclerosis increase the pressure on the intracranial artery wall, thereby elevating the risk of arterial tissue remodeling and intracranial aneurysm formation^8^. However, the relationship between atherosclerosis and the development of intracranial aneurysms remains complex and is not fully understood, with ongoing debate about whether they share common risk factors or whether there is a direct causal link between them.

The potential comorbidity and causal relationship between atherosclerosis and intracranial aneurysms have not been definitively established. This study aimed to investigate the prevalence of intracranial aneurysms and atherosclerosis based on a real-world cross-sectional study and to utilize Mendelian randomization to explore the possible causal relationships and common causative factors between these two diseases. Finally, the common pathogenic biomarkers identified through Mendelian randomization were validated in clinical samples.

## Methods

### 1. Study design

This study explores the potential association between atherosclerosis and intracranial aneurysms, divided into four sections based on the progression of the analysis: real-world cross-sectional study, Mendelian randomization analysis, plasma validation, and in situ specimen validation. The detailed arrangement is shown in Figure 1. This study adhered to the guidelines outlined in the Declaration of Helsinki and received approval from the Ethics Committee of Beijing Tiantan Hospital, which is affiliated with Capital Medical University (Ethics approval number: KY2022-051-02). Prior to enrollment, all participants provided informed consent for the collection of clinical information and imaging data.

**Figure 1.**
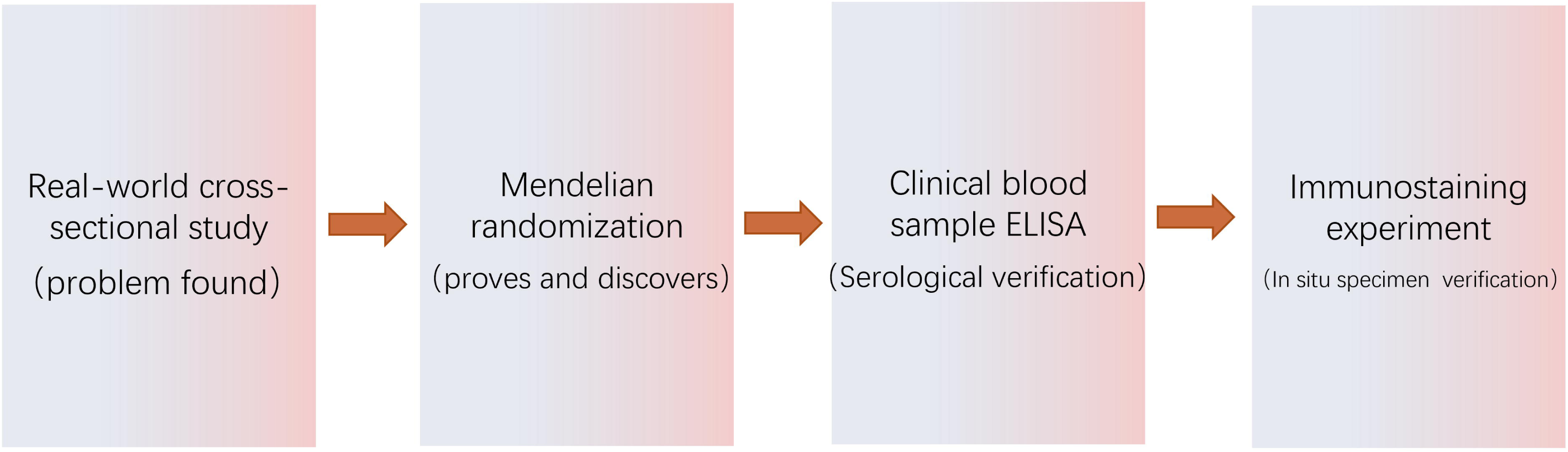
Research arrangement.

### 2. Real-world cross-sectional study

The study population consisted of patients who underwent head Computed tomography angiography (CTA) or MRA examinations at Beijing Tiantan Hospital between January 1, 2023, and August 31, 2023. The inclusion criteria were as follows: (1) Head CTA or MRA examination with a diagnosis including the term ‘plaque’. (2) Head CTA or MRA examination with a description including the terms ‘stenosis’ or ‘occlusion’. Patients who had undergone multiple examinations were excluded, and only the results of their first examination were considered. The final sample size for the cross-sectional study was 13,739, and data on sex, age, and intracranial aneurysm incidence were collected. Intracranial aneurysm diagnosis was based on head CTA or MRA examinations that included the term ‘aneurysm’.

### 3. Mendelian Randomization analysis

We performed two-sample MR to investigate three sets of associations: (1) the association between genetic susceptibility to atherosclerosis and intracranial aneurysms; (2) the association between genetic susceptibility to atherosclerosis and intracranial aneurysms with circulating matrix metalloproteinase family levels; and (3) the genetically predicted association of circulating matrix metalloproteinase family levels with the risk of atherosclerosis and intracranial aneurysms.

We analyzed the causal relationship between atherosclerosis and intracranial aneurysms, considering the various subtypes of each disease. Intracranial aneurysms were classified as either unruptured or ruptured, while atherosclerosis was categorized into four subtypes based on location: cerebral atherosclerosis, coronary atherosclerosis, peripheral atherosclerosis and other atherosclerosis. Among them, peripheral atherosclerosis refers to atherosclerosis of the limbs. Other atherosclerosis refers to atherosclerosis occurring in all locations except the cerebral arteries, coronary arteries, and peripheral arteries of the limbs.

To investigate the potential role of plasma MMPs in the relationship between these two diseases, we selected 12 MMPs for analysis using a two-way MR approach. Our objective was to identify plasma MMPs associated with or induced by intracranial aneurysms and atherosclerosis and to evaluate their potential role in mediating the causal relationship between these two diseases.

### 4. Instrument Selection

The sources of genetic instrumental variables are presented in Table 2. We performed two-sample MR analysis and two-way MR analysis using genetic instrumental variables with p values less than 5×10^-9 for each exposure. If the genetic instrumental variables could not be identified with this p value, we adjusted the threshold to less than 5×10^-6. We excluded associated SNPs (r^2 > 0.1) by retaining the SNP with the smallest p value associated with the SNP exposure association. To avoid weak instrument bias, we included only SNPs with an F-statistic greater than 10^9^.

### 5. Sensitivity analysis

We performed sensitivity analyses using the MR-Egger, weighted median, and MR Pleiotropic Residual Sum and Outlier (MR-PRESSO) methods to evaluate robustness and assess horizontal pleiotropic effects when more than two instrumental variables were available. MRLEgger analysis estimated the pleiotropic effect of the instrumental variable, with nonzero intercepts indicating bias in the inverse variance weighted (IVW) estimates. The weighted median approach provides a causal estimate by considering the nullity of multiple genetic variants or the presence of pleiotropic effects. MR-PRESSO used a global test to detect horizontal pleiotropy and, if necessary, corrected for potential pleiotropic outliers by removing them^10^. The heterogeneity of the IVW estimates was evaluated using Cochran’s Q test^11^.

We performed bidirectional MR analyses to further explore the directionality of potential reciprocal relationships between diseases and between biomarkers and diseases. To correct for multiple testing, we used the Bonferroni method. In the MR analysis of the association between atherosclerosis and intracranial aneurysms, we used a p value threshold of 0.05/8 = 6.25×10^-3. In the two-way MR analysis of MMPs levels and diseases of interest, we considered 72 unique associations with a p value threshold of 0.05/72 = 6.94×10^-4. P values below the threshold after multiple comparisons were considered strong evidence of association, while those between the threshold and 0.05 were considered suggestive of associations. All analyses were two-sided and performed using the TwoSampleMR (version 1.0.3) and MRPRESSO (version 6.3) packages in R software (version 4.2.1). The report adheres to the STROBE-MR statement.

### 6. Plasma ELISA and in situ specimen validation

Detailed materials and methods for serum ELISA and in situ specimen validation are provided in the supplementary materials.

### 7. Statistical analysis

All analyses were performed using SPSS (version 25) and R (version 4.1.2). Categorical variables are presented as counts (percentages), and comparisons of proportions were conducted using the Pearson chi-square test. Continuous variables are described as either the mean ± standard deviation or median and interquartile range, depending on their distribution, which was assessed by the KolmogorovLSmirnov test. All the statistical tests were two-sided.

## Results

### Cross-sectional study

This cross-sectional study involved a total of 13,739 participants, comprising 2,946 patients with intracranial aneurysms and 1,180 patients with cerebral artery atherosclerosis. The average age of the participants was 59 years, with a male population of 48%. Age did not differ significantly between patients with and without intracranial aneurysms. However, a greater proportion of patients with aneurysms were female (60.692%) than male patients (39.308%). The sex distribution varied between the groups. Additionally, patients with intracranial aneurysms exhibited a greater prevalence of cerebral artery atherosclerosis than did those without intracranial aneurysms (14.155% vs. 7.069%, P<0.001). The original data of the cross-sectional studies are shown in eTable 1, and the baseline data are shown in Table 1. Even after adjusting for age and sex, multivariate regression analysis indicated that cerebral artery atherosclerosis remained an association with intracranial aneurysm (OR=2.439, 95% CI 2.139-2.781; P<0.001).

**Table 1.**
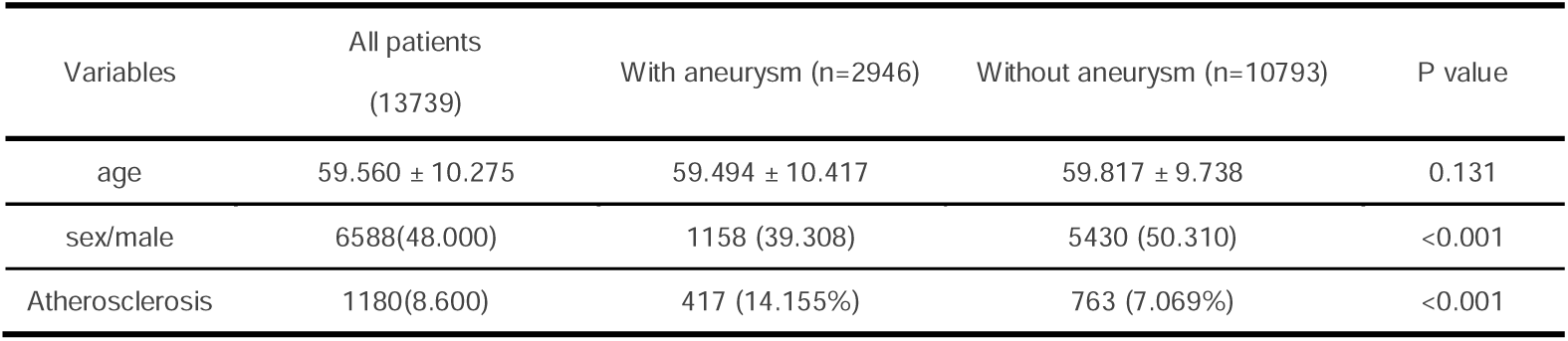
Cross-sectional study baseline table.

| Variables | All patients<br>(13739) | With aneurysm (n=2946) | Without aneurysm (n=10793) | P value |
| --- | --- | --- | --- | --- |
| age | 59.560 ± 10.275 | 59.494 ± 10.417 | 59.817 ± 9.738 | 0.131 |
| sex/male | 6588(48.000) | 1158 (39.308) | 5430 (50.310) | <0.001 |
| Atherosclerosis | 1180(8.600) | 417 (14.155%) | 763 (7.069%) | <0.001 |

**Table 2.** Sources of Mendelian randomization genetic instruments.

| Trials | Cases (N) | Controls (N) | Sample size (N) | Year of publication | No. of SNPs | Population | Source |
| --- | --- | --- | --- | --- | --- | --- | --- |
| Cerebral aneurysm,<br>nonruptured | 992 | 217,800 | 218,792 | 2021 | 16,380,466 | European | FinnGen GWAS |
| Cerebral aneurysm, SAH | 2,127 | 216,665 | 218,792 | 2021 | 16,380,466 | European | FinnGen GWAS |
| Coronary atherosclerosis | 23,363 | 187,840 | 211,203 | 2021 | 16,380,447 | European | FinnGen GWAS |
| Cerebral atherosclerosis | 104 | 218,688 | 218,792 | 2021 | 16,380,466 | European | FinnGen GWAS |
| Peripheral atherosclerosis | 6,631 | 162,201 | 168,832 | 2021 | 16,380,247 | European | FinnGen GWAS |
| Matrix metalloproteinase-1 | / | / | 21,758 | 2020 | 12,705,782 | European | Folkersen L et al. |
| Matrix metalloproteinase-2 | / | / | 1,000 | 2019 | 501,428 | European | Suhre K et al. |
| Matrix metalloproteinase-3 | / | / | 21,758 | 2020 | 13,057,986 | European | Folkersen L et al. |
| Matrix metalloproteinase-7 | / | / | 21,758 | 2020 | 12,938,892 | European | Folkersen L et al. |
| Matrix metalloproteinase-8 | / | / | 1,000 | 2019 | 501,428 | European | Suhre K et al. |
| Matrix metalloproteinase-9 | / | / | 3,301 | 2018 | 10,534,735 | European | Sun BB et al. |
| Matrix metalloproteinase-10 | / | / | 21,758 | 2020 | 12,718,221 | European | Folkersen L et al. |
| Matrix metalloproteinase-12 | / | / | 21,758 | 2020 | 13,100,092 | European | Folkersen L et al. |
| Matrix metalloproteinase-13 | / | / | 1000 | 2019 | 501,428 | European | Suhre K et al. |
| Matrix metalloproteinase-14 | / | / | 3,301 | 2018 | 10,534,735 | European | Sun BB et al. |
| Matrix metalloproteinase-16 | / | / | 3,301 | 2018 | 10,534,735 | European | Sun BB et al. |
| Matrix metalloproteinase-17 | / | / | 3,301 | 2018 | 10,534,735 | European | Sun BB et al. |

**GWAS, genome-wide association study**

### Two-sample Mendelian randomization analysis (atherosclerosis and intracranial aneurysm)

Using double-sample MR analysis, we investigated the genetically predicted relationship between atherosclerosis and intracranial aneurysm, with atherosclerosis as the exposure and intracranial aneurysm as the outcome. Figure 2 presents the results of eight consecutive analyses with P values less than 0.05. After adjusting for multiple comparisons, we found that genetically predicted peripheral atherosclerosis increased the risk of intracranial aneurysms, including unruptured intracranial aneurysms (OR_IVW_ = 1.711; 95% CI 1.208-2.423; P_IVW_ = 0.002) and ruptured intracranial aneurysms (OR_IVW_ = 1.533; 95% CI 1.190-1.975; P_IVW_ = 9.41×10^-4). MR-IVW did not present heterogeneity. The MR-PRESSO test showed no horizontal pleiotropy. This causal association persisted after Bonferroni multiple correction. In addition, when considering intracranial aneurysm as an exposure factor and atherosclerosis as an outcome, results indicate that ruptured intracranial aneurysms are a risk factor for cerebral atherosclerosis (OR_MR-Egger_ = 4.203; 95% CI 1.293-13.658; P_MR-Egger_ = 0.034). However, the p-value did not pass multiple testing corrections, and the results exhibit pleiotropy (Pleiotropy P Value <0.05). The summary results and sensitivity analyses of the two-sample analysis are shown in eTable 6. The genetic instruments used for dual-sample MR analysis are shown in eTables 2 and 3.

**Figure 2.**
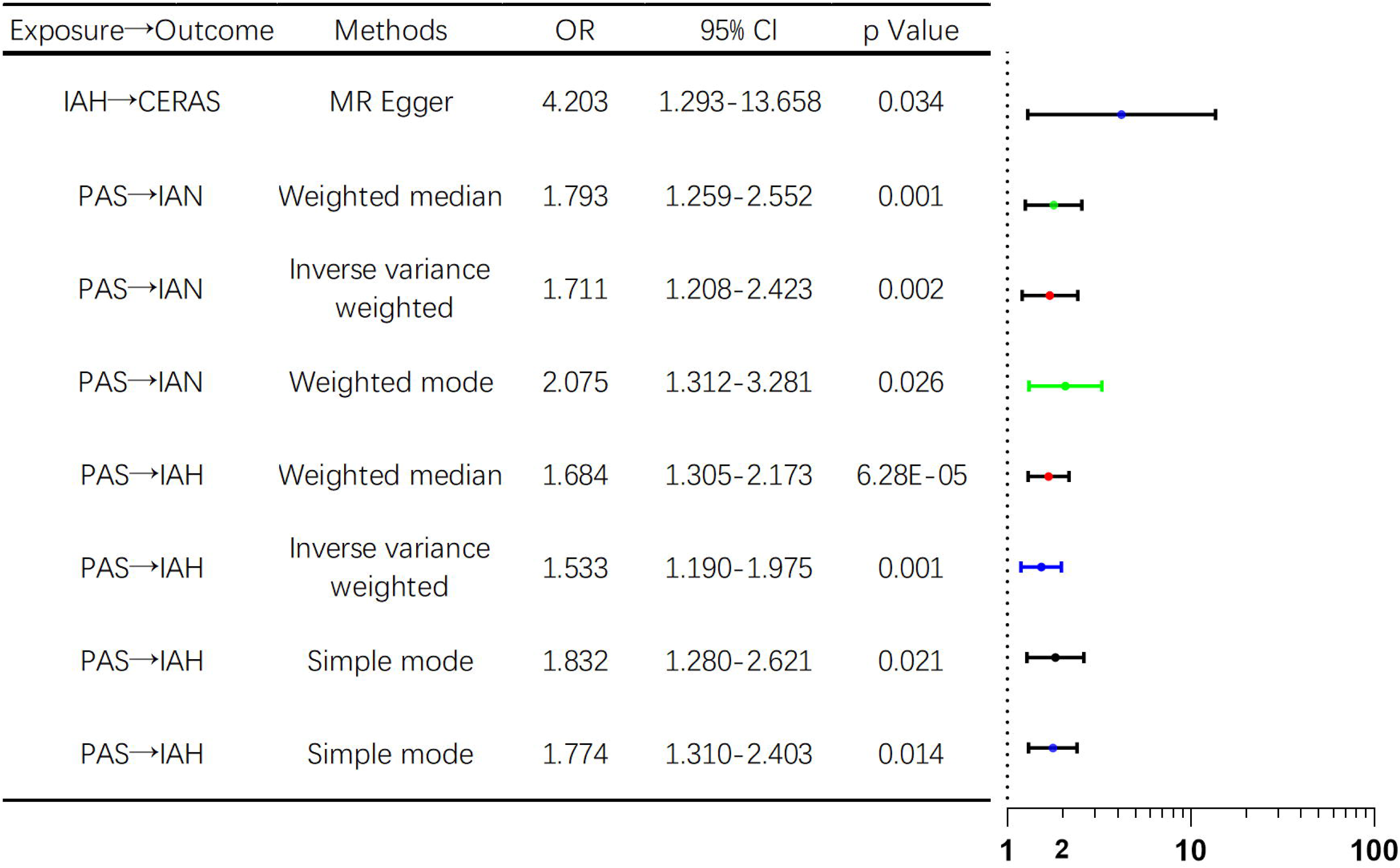
Two-way MR analysis of the associations between atherosclerosis and intracranial aneurysms (P < 0.05). CERAS, cerebral atherosclerosis; PAS, peripheral atherosclerosis; IAH, ruptured intracranial aneurysm; IAN, unruptured intracranial aneurysm.

### Bidirectional Mendelian Randomization (Atherosclerosis/Intracranial Aneurysm and Matrix Metalloproteinases)

After establishing the causal relationship between atherosclerosis and intracranial aneurysms, we performed a two-way MR analysis to investigate the genetic predictive association between matrix metalloproteinase levels and the target diseases and to determine whether matrix metalloproteinases act as a cause or a result of the diseases. Figure 3 displays the results with P values less than 0.05. Our analysis revealed a genetically elevated risk of peripheral atherosclerosis, resulting in elevated circulating MMP10 levels (β_IVW_ = 0.087; 95% CI 0.007 to 0.167; P_IVW_ = 0.033), MMP12 levels (β_IVW_ = 0.083; 95% CI 0.022 to 0.145; P_IVW_ = 0.008) and MMP13 levels (β_IVW_ = 0.399; 95% CI 0.063 to 0.735; P_IVW_ = 0.020). A genetically predicted elevated risk for coronary atherosclerosis was associated with increased circulating MMP14 levels (β_IVW_ = 0.141; 95% CI 0.050 to 0.232; P_IVW_ = 0.002). Unruptured brain aneurysms reduce circulating MMP2 levels (β_MR Egger_ = -0.425; 95% CI -0.788 to -0.062; P_MR Egger_ = 0.041) but increase MMP13 levels (β_IVW_ = 0.116; 95% CI 0.001 to 0.230; P_IVW_ = 0.048). Other atherosclerotic risks increase MMP10 levels (β_IVW_ = 0.078; 95% CI 0.011 to 0.145; P_IVW_ = 0.022). MR-IVW and MR Egger did not present heterogeneity. The MR-PRESSO test showed no horizontal pleiotropy.

**Figure 3.**
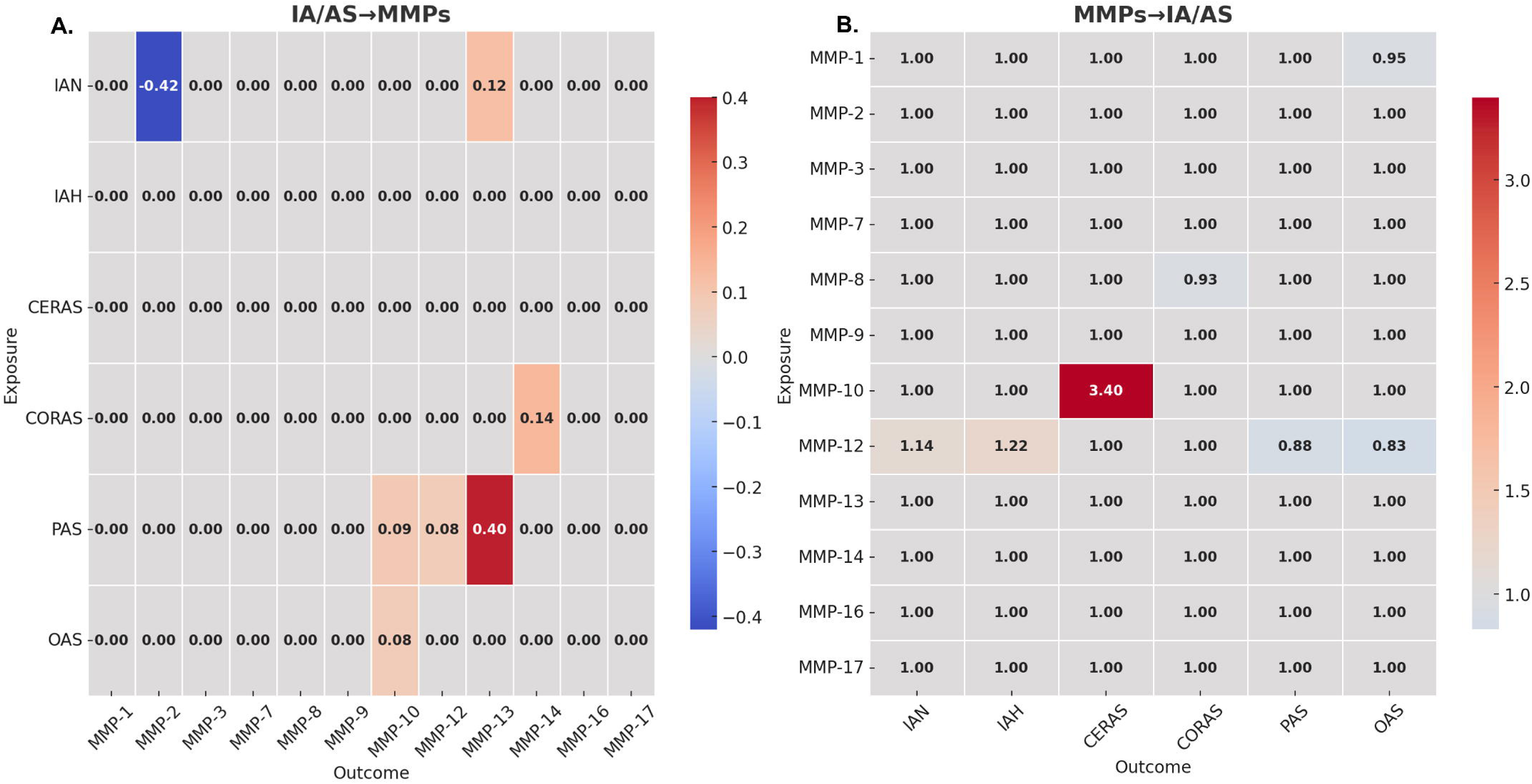
Two-way MR analysis of the associations between circulating MMPs levels and atherosclerosis and intracranial aneurysms (P < 0.05). CORAS, coronary atherosclerosis; CERAS, cerebral atherosclerosis; PAS, peripheral atherosclerosis; OAS, other atherosclerosis; IAH, ruptured intracranial aneurysm; IAN, unruptured intracranial aneurysm.

Next, we analyzed the impact of circulating MMPs levels on genetic susceptibility to the two target diseases. After investigating 12 MMPs, we discovered that higher circulating MMP12 levels were correlated with increased risks of unruptured intracranial aneurysms (OR_IVW_ =1.136; 95% CI 1.031–1.252; P_IVW_ =0.010) and ruptured intracranial arteries (OR_simple mode_ = 1.217; 95% CI 1.016–1.458; P_simple mode_ = 0.038), while they were associated with a reduced risk of peripheral atherosclerosis (OR_IVW_ =0.883; 95% CI 0.846–0.922; P_IVW_ =1.47×10^-8) and other atherosclerosis (OR_IVW_ =0.893; 95% CI 0.855–0.933; P_IVW_ =4.62×10^-7). Additionally, elevated levels of circulating MMP10 were linked to a heightened risk of cerebral atherosclerosis (OR_MR Egger_ =3.398; 95% CI 1.183–9.757; P_MR Egger_ =0.033), while higher levels of circulating MMP8 were related to a decreased risk of coronary atherosclerosis (OR_MR Egger_ =0.933; 95% CI 0.886–0.983; P_MR Egger_ =0.021). And higher levels of circulating MMP1 were related to a decreased risk of other atherosclerosis (OR_IVW_ =0.948; 95% CI 0.899–0.999; P_IVW_ =0.045). MR-IVW and MR Egger did not present heterogeneity. The MR-PRESSO test showed no horizontal pleiotropy. The summary results and sensitivity analyses of the two-sample analysis are shown in eTable 7 and 8. The genetic instruments used for dual-sample MR analysis are shown in eTables 4 and 5.

Two-way MR analysis of plasma matrix proteases and intracranial aneurysm and atherosclerotic disease, as described earlier, revealed a reciprocal correlation between MMP12 levels and the risk of peripheral atherosclerosis. Although the above causal relationships disappear after Bonferroni multiple correction, they still have certain reference values.

### Plasma ELISA verification

To verify the common key pathogenic factor of atherosclerosis and intracranial aneurysms identified through MR, specifically the role of plasma MMP12 in these conditions, we collected samples from patients with carotid atherosclerosis and intracranial aneurysms, as well as from a control group of healthy participants. Plasma samples were collected and measured using ELISA. The clinical baseline characteristics and plasma MMP12 levels for carotid artery stenosis patients, intracranial aneurysm patients, and normal controls are presented in Table 3. Compared to healthy controls, patients with aneurysms and carotid stenosis were older and had greater systolic blood pressure. Plasma MMP12 levels were significantly greater in patients with carotid atherosclerosis than in controls. Although plasma MMP12 levels in patients with intracranial aneurysms were greater than those in the control group, the difference was not statistically significant. To further control for confounding factors, we conducted a multivariate logistic regression analysis. The results indicated that plasma MMP12 levels (CAS: OR = 2.284, 95% CI 1.357-4.638, P = 0.007; IA: OR = 1.557, 95% CI 1.067-2.426, P = 0.032), age group (CAS: OR = 21.473, 95% CI 3.622-199.404, P = 0.002; IA: OR = 5.933, 95% CI 1.711-23.742, P = 0.007), and systolic blood pressure (CAS: OR = 1.129, 95% CI 1.022-1.274, P = 0.028; IA: OR = 1.099, 95% CI 1.033-1.185, P = 0.006) are risk factors for carotid artery stenosis and intracranial aneurysms. The regression analysis results are shown in Table 4.

**Table 3.** Baseline table of patients with atherosclerosis and intracranial aneurysm serum.

| Disease | Variables | All participants (n=79) | HC (n=35) | IA (n=44) | P value | Disease | Variables | All participants (n=73) | HC (n=35) | CAS (n=38) | P value |
| --- | --- | --- | --- | --- | --- | --- | --- | --- | --- | --- | --- |
| intracranial<br>aneurysm | age>40 | 53(67.089) | 16(45.714) | 37(84.091) | <0.001 | Carotid<br>artery<br>stenosis | age>50 | 45(61.644) | 10(28.571) | 35(92.105) | <0.001 |
|  | sex/female | 42(53.165) | 17(48.571) | 25(56.818) | 0.466 |  | sex/female | 21(28.767) | 17(48.571) | 4(10.526) | <0.001 |
|  | BMI, kg/m2 | 24.526±3.991 | 23.388±3.732 | 25.431±3.958 | 0.024 |  | BMI, kg/m2 | 23.875±3.538 | 23.388±3.732 | 24.324±3.287 | 0.265 |
|  | SBP/mmHg | 130.000[124.000,138.000] | 126.000[115.000,130.000] | 135.000[130.000,140.000] | <0.001 |  | SBP/mmHg | 130.000[122.000,136.000] | 126.000[115.000,130.000] | 133.000[130.000,138.000] | 0.001 |
|  | DBP/mmHg | 80.709±8.748 | 78.886±7.678 | 82.159±9.261 | 0.101 |  | DBP/mmHg | 80.000[75.000,83.000] | 79.000[75.000,81.000] | 80.000[76.000,85.000] | 0.516 |
|  | Heart Rate | 78.101±9.703 | 76.657±9.905 | 79.250±9.381 | 0.244 |  | Heart Rate | 78.000[72.000,80.000] | 76.000[70.000,80.000] | 80.000[76.000,82.000] | 0.049 |
|  | **Medical history** |  |  |  |  |  | **Medical history** |  |  |  |  |
|  | coronary heart<br>disease | 4(5.063) | 0(0.000) | 4(9.091) | NA |  | coronary heart<br>disease | 6(8.219) | 0(0.000) | 6(15.789) | NA |
|  | Hyperlipidemia | 3(3.797) | 0(0.000) | 3(6.818) | NA |  | Hyperlipidemia | 7(9.589) | 0(0.000) | 7(18.421) | NA |
|  | diabetes | 5(6.329) | 0(0.000) | 5(11.364) | NA |  | diabetes | 14(19.178) | 0(0.000) | 14(36.842) | NA |
|  | hypertension | 26(32.911) | 0(0.000) | 26(59.091) | NA |  | hypertension | 27(36.986) | 0(0.000) | 27(71.053) | NA |
|  | Cigarette smoking | 7(8.861) | 0(0.000) | 7(15.909) | NA |  | Cigarette smoking | 19(26.027) | 0(0.000) | 19(50.000) | NA |
|  | Alcohol drinking | 7(8.861) | 0(0.000) | 7(15.909) | NA |  | Alcohol drinking | 16(21.918) | 0(0.000) | 16(42.105) | NA |
|  | **MMP12** | 5.769[4.908,6.508] | 5.182[3.816,6.562] | 5.969[5.285,6.508] | 0.056 |  | **MMP12** | 6.439±2.041 | 5.382±1.820 | 7.413±1.725 | <0.001 |

**HC, healthy control; IA, intracranial aneurysm; CAS, carotid artery stenosis; BMI, body mass index; SBP, systolic blood pressure; DBP, diastolic blood pressure.**

**Table 4.** Logistic regression analysis of the associations between serum MMP12 levels and atherosclerosis and intracranial aneurysms.

| Disease | Variables | Estimate | SE | Z | P value | Odds Ratio | Lower | Upper |
| --- | --- | --- | --- | --- | --- | --- | --- | --- |
| intracranial aneurysm | age>40 | 1.781 | 0.661 | 2.695 | 0.007 | 5.933 | 1.711 | 23.742 |
|  | sex/female | 0.39 | 0.629 | 0.62 | 0.535 | 1.477 | 0.429 | 5.219 |
|  | BMI, kg/m2 | 0.122 | 0.076 | 1.599 | 0.11 | 1.129 | 0.979 | 1.325 |
|  | SBP/mmHg | 0.095 | 0.035 | 2.729 | 0.006 | 1.099 | 1.033 | 1.185 |
|  | DBP/mmHg | -0.035 | 0.042 | -0.823 | 0.41 | 0.966 | 0.886 | 1.048 |
|  | Heart Rate | 0.041 | 0.034 | 1.198 | 0.231 | 1.042 | 0.976 | 1.119 |
|  | MMP12 | 0.443 | 0.206 | 2.144 | 0.032 | 1.557 | 1.067 | 2.426 |
| Carotid artery stenosis | age>50 | 3.067 | 0.998 | 3.074 | 0.002 | 21.473 | 3.622 | 199.404 |
|  | sex/female | -1.327 | 0.997 | -1.33 | 0.183 | 0.265 | 0.032 | 1.778 |
|  | BMI, kg/m2 | -0.028 | 0.146 | -0.192 | 0.847 | 0.972 | 0.732 | 1.306 |
|  | SBP/mmHg | 0.121 | 0.055 | 2.196 | 0.028 | 1.129 | 1.022 | 1.274 |
|  | DBP/mmHg | -0.033 | 0.069 | -0.483 | 0.629 | 0.967 | 0.836 | 1.105 |
|  | Heart Rate | 0.08 | 0.053 | 1.518 | 0.129 | 1.084 | 0.98 | 1.213 |
|  | MMP12 | 0.826 | 0.306 | 2.702 | 0.007 | 2.284 | 1.357 | 4.638 |

BMI, body mass index; SBP, systolic blood pressure; DBP, diastolic blood pressure.

### MMP12 immunohistochemical staining

To investigate the local presence of MMP12 in atherosclerotic and aneurysmal tissues, we conducted immunohistochemical staining of MMP12 in normal fibrous tissue distal to human carotid plaques, carotid plaques, aneurysms, and normal carotid plaques in model mice. Additionally, mouse cerebral blood vessels were fluorescently stained for MMP12. Our findings revealed clear MMP12 positivity in carotid plaque tissue, while no positive staining was observed in distal normal fibrous tissue. The plaque staining results are shown in Figure 4A. In model mice, MMP12 was found to be distributed in the elastin layer of aneurysm tissue but not in the cerebral blood vessels of normal mice. The staining results of intracranial aneurysms are shown in Figure 4B. These results indicate the widespread presence of MMP12 in the local tissues of arterial plaques and aneurysms, suggesting its potential involvement in the pathogenesis of atherosclerosis and aneurysms and confirming our initial hypothesis.

**Figure 4.**
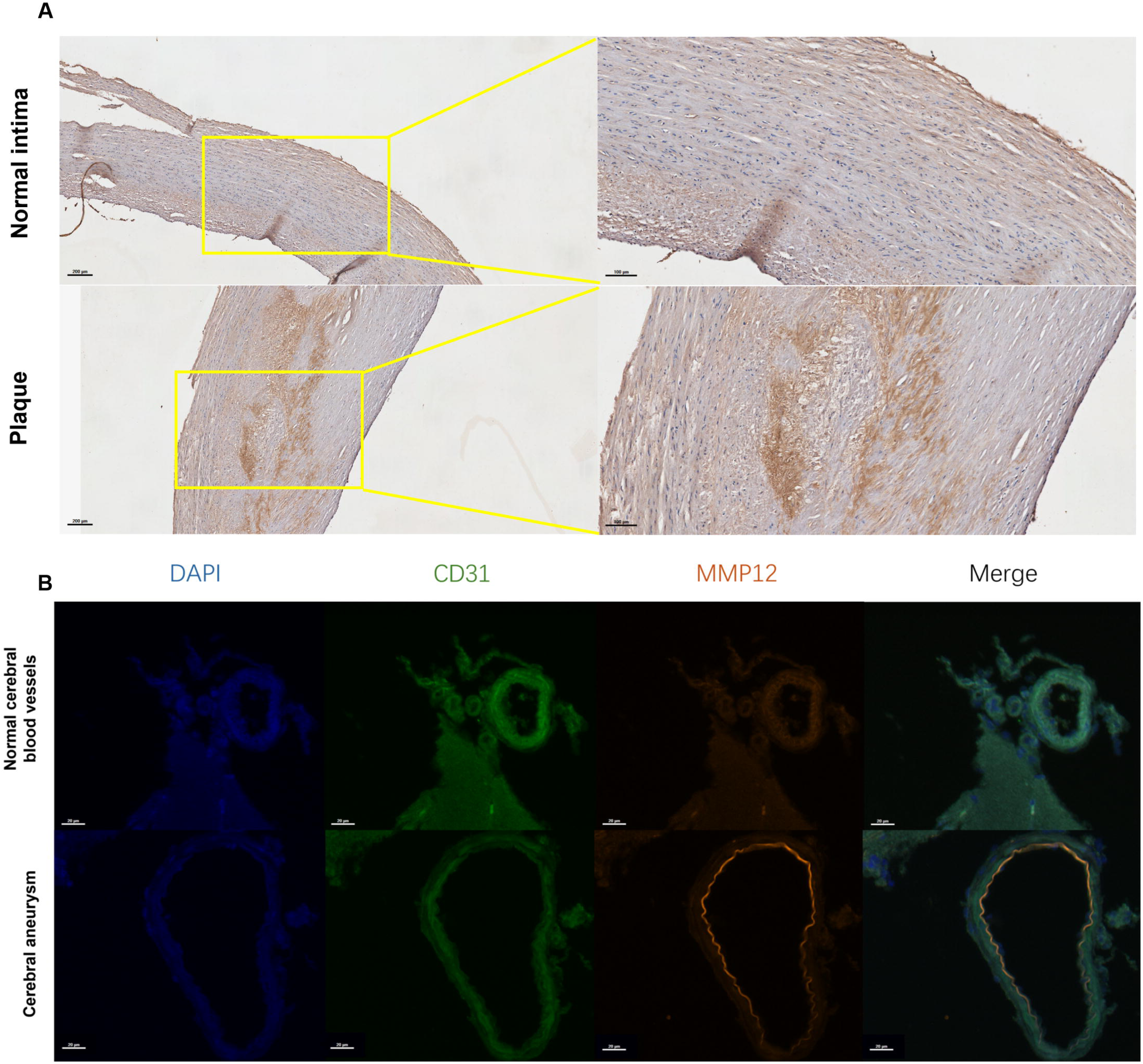
A. Immunohistochemical staining of MMP12 in the necrotic area in the core of the carotid plaque and normal fibrous tissue in the distal region; B. MMP12 immunofluorescence staining of cerebral blood vessels in intracranial aneurysm model mice and normal mice.

## Discussion

The cross-sectional investigation revealed that patients with intracranial aneurysms have a greater incidence of atherosclerosis. Two-sample MR analysis indicated an association between peripheral atherosclerosis and the risk of intracranial aneurysms. In view of the role of matrix metalloproteinases in atherosclerosis and aneurysms in previous literature^12,13^, we performed a bidirectional MR analysis on the relationships between MMPs and both atherosclerosis and intracranial aneurysms. Mendelian randomization results indicate that an increased prevalence of peripheral arterial atherosclerosis is associated with elevated circulating MMP12 levels. Additionally, elevated circulating MMP12 is associated with an increased risk of intracranial aneurysm. The results of subsequent ELISA and specimen validation were consistent with the findings from Mendelian randomization. Overall, our research confirms a causal link between atherosclerosis and the development of intracranial aneurysms, identifying and validating MMP12 as a relevant biomarker.

The relationship between atherosclerosis and aneurysm development is complex and multifaceted. Both conditions share common risk factors, including smoking, hypertension, and hypercholesterolemia^14^. Although atherosclerotic plaques are often found in aneurysmal walls^7^, it remains unclear whether atherosclerosis directly causes aneurysms. One theory suggests that aneurysms are a pathological response to atherosclerosis^15^. The most compelling arguments focus on arterial remodeling, with extensive in vitro, animal, and histological data indicating that compensatory changes occur in response to lumen narrowing and shear stress, promoting arterial expansion and potentially leading to severe medial thinning^16^. Proinflammatory cytokines from atherosclerotic or thrombotic inflammation combined with elastin degradation stimulated by medial proteolysis may trigger a chronic inflammatory response^17^. Another theory posits that atherosclerosis and aneurysm development are independent but parallel processes influenced by common environmental and genetic risk factors. A third perspective suggests that either condition can precede or stimulate the other^18^. Johnsen et al. analyzed data from the Tromsø Study and found no consistent doseLresponse relationship between the extent of atherosclerosis and abdominal aortic aneurysm size, implying simultaneous development rather than a direct causative link^19^. However, regional factors such as hemodynamics influence severity, suggesting that the study’s cross-sectional design limits its ability to assess disease onset relationships prospectively.

Several studies have also analyzed the association between atherosclerosis and the rupture of intracranial aneurysms. The results indicate that the quantity and severity of cerebral arterial atherosclerosis are significantly correlated with the risk of aneurysm rupture^20^. Additionally, the intima thickness of carotid plaques may serve as a potential predictor for aneurysm rupture^21^. Therefore, it can be inferred that atherosclerosis may be associated with the progression of intracranial aneurysms.

In summary, previous research has predominantly relied on retrospective analyses exploring indirect associations between these two conditions, resulting in ambiguous and uncertain conclusions. No previous studies have used MR to analyze their relationships, making our research the first to provide a clear answer on causal associations from a genetic perspective using this method. Furthermore, to explore the mechanisms underlying atherosclerosis-induced intracranial aneurysms, we identified MMP12 as a key mediator through MR and validated its role through serological and in situ specimen analyses. MMP12, a zinc-dependent endopeptidase involved in extracellular matrix (ECM) degradation, is secreted primarily by macrophages and is crucial for tissue remodeling, inflammation, and various pathological conditions^22,23^. Notably, MMP12 is significantly upregulated in atherosclerotic lesions^24,25^. During the development of atherosclerosis, damage to the vascular wall triggers an inflammatory response, leading to the aggregation of inflammatory cells such as macrophages, which secrete MMP12^26^. Additionally, in atherosclerotic lesions, activated vascular smooth muscle cells also produce MMP12, resulting in elevated MMP12 levels^27^. Elevated MMP12 levels have been linked to abdominal aortic aneurysms, promoting ECM degradation and arterial wall weakening^28^. Studies using MMP12 knockout mice demonstrated that MMP12 deficiency significantly impaired aneurysm growth and reduced elastin fiber degradation, underscoring its critical role in aneurysm development and progression^29^. Although the direct role of MMP12 in intracranial aneurysms has not been well established, similar ECM degradation and inflammatory mechanisms may be involved. The ability of MMP12 to degrade elastin and other structural proteins may weaken arterial walls, potentially facilitating intracranial aneurysm formation and rupture^22^. While previous research has reported the roles of MMP12 in atherosclerosis and aneurysms, no study has linked MMP12 to both conditions. Our study confirmed that atherosclerosis induces elevated plasma MMP12, which promotes the development of intracranial aneurysms.

Our study has limitations. First, the cross-sectional data were derived from a single center and relied on keyword searches within the imaging system, potentially introducing selection bias. Additionally, the baseline information included only age and sex and lacked information on other comorbidities and risk factors, such as smoking and drinking, preventing us from excluding the influence of these traditional risk factors on the prevalence of atherosclerosis and intracranial aneurysms. Second, in our analysis of the association between MMPs and the two diseases, the causal relationship derived from our MR study lost significance after Bonferroni correction, suggesting a weaker association and the potential existence of more meaningful biomarkers. However, this result was validated through serological and in situ specimen experiments, indicating that the identified biomarker still holds reference value. Third, all MR analyses were based on individuals of European descent, limiting the applicability of the results to Asian populations. Although the serological and atherosclerotic plaque data were derived from Asian populations and aligned with the MR findings, further studies with larger sample sizes are needed to confirm generalizability. Fourth, this study did not conduct prospective analyses of the real-world association between atherosclerosis and intracranial aneurysms, necessitating further large-scale cohort studies to observe incidence rates. Finally, in our Mendelian randomization analysis, we observed a significant promoting effect of peripheral atherosclerosis on intracranial aneurysm formation. However, no significant association was found between intracranial atherosclerosis and intracranial aneurysms. It is important to note that the results of Mendelian randomization are intended to provide clues, and they are specific to the data used in this study. Negative results only suggest that no significant genetic association was found, and do not imply that they do not play a role in disease development in the real world. Therefore, further research is needed to explore the impact of atherosclerosis at different sites on intracranial aneurysms.

## Conclusion

This study utilized a large-scale cross-sectional dataset to investigate the relationship between atherosclerosis and the incidence of intracranial aneurysms. Through MR analysis, we explored causal relationships and identified intermediate biomarkers, ultimately validating these biomarkers through serological and in situ specimen immunostaining experiments. Our research elucidates the causal link between atherosclerosis and intracranial aneurysms by identifying and verifying MMP12 as a critical biomarker, thereby providing new methods and evidence to explain the association between these two conditions.

## List of abbreviations

MMP: matrix metalloproteinase
MR: Mendelian Randomization
MRA: magnetic resonance angiography
AWE: aneurysm wall enhancement
CTA: Computed tomography angiography
GWAS: genome-wide association study
SNPs: single nucleotide polymorphisms
MR-PRESSO: MR pleiotropic residual sum and outlier
IVW: inverse-variance weighted
ECM: extracellular matrix
CAS: carotid atherosclerosis
IA: intracranial aneurysm
IAN: unruptured intracranial aneurysm
IAH: ruptured intracranial aneurysm
CERAS: cerebral atherosclerosis
CORAS: coronary atherosclerosis
PAS: peripheral atherosclerosis
OAS: Other atherosclerosis

## Supporting information

eTables

## Data Availability

The raw data supporting the conclusions of this article will be made available by the authors without undue reservation. All data used for Mendelian randomization analyses are publicly available from the respective GWAS.

https://1drv.ms/x/c/5ab744d869ae75bd/EcCsnlPF4GBGm39I_wipcY8Bd0GFI-Yc63tUKYSGC0Av_A?e=dhqVeV

## Declarations

### Ethics statement

The studies involving human participants were reviewed and approved by Beijing Tiantan Hospital, Capital Medical University (Ethics approval number: KY2022-051-02). The patients/participants provided written informed consent to participate in this study.

All the animals were acclimated under standard laboratory conditions (ventilated room, 25±1 °C, 60 ± 5 % humidity, 12 h light/dark cycle) and had free access to standard water and food(SYXK-2024-0003). All procedures were conducted in accordance with the “ Guiding Principles in the Care and Use of Animals” (China) and were approved by theLaboratory Animal Ethics Committee of Beijing Neurosurgical Institute (BNI202201011).

### Consent for publication

Not applicable.

### Authors’ contributions

Dong Zhang and Peicong Ge conceived the idea; Wei Liu conducted the analyses and wrote the paper; Zhaoxu Zheng developed the animal model and provided intracranial aneurysm specimens. Chenglong Liu, Yuanren Zhai, Shuang Wang and Liangran Huang provided the data; all authors contributed to the revisions.

