## supplementary figure for "Atherosclerosis, Intracranial Aneurysms, and Intermediate Biomarkers: Real-World Observational and Mendelian Randomization Research"

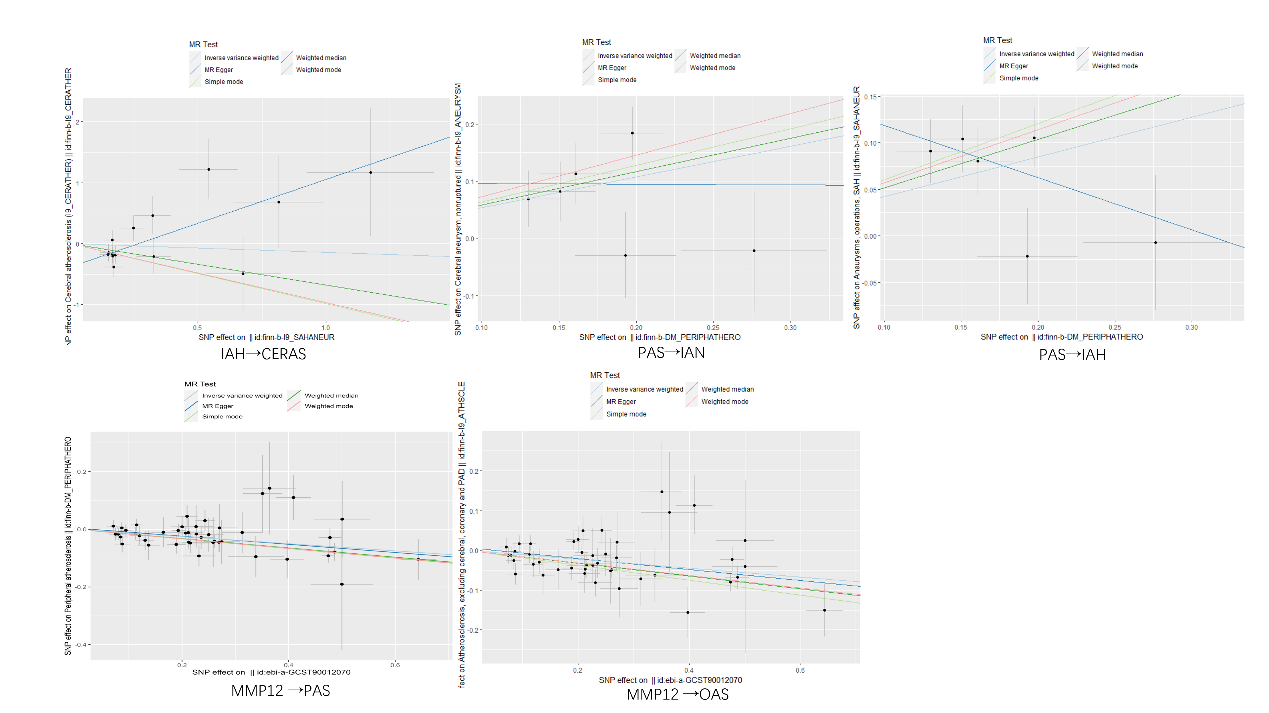

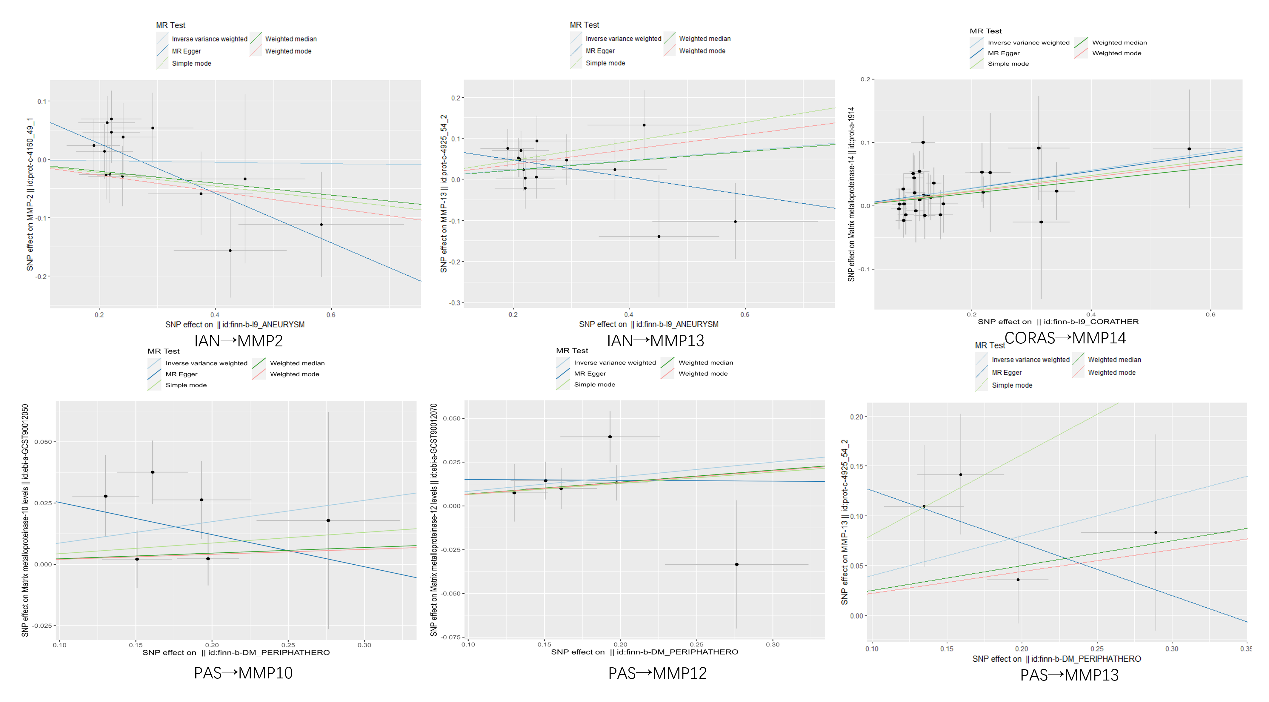

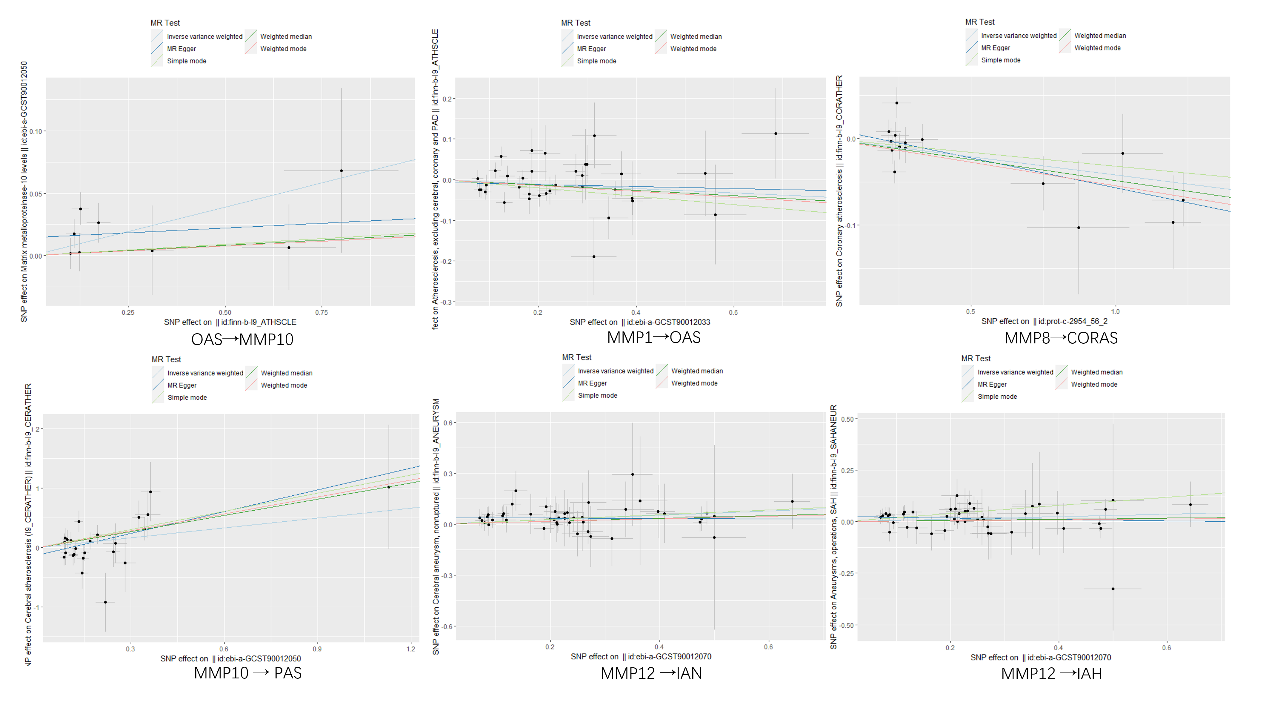

**Supplementary Figures 1-3.** Scatter plots of results with p-values less than 0.05 in Mendelian randomization analysis.

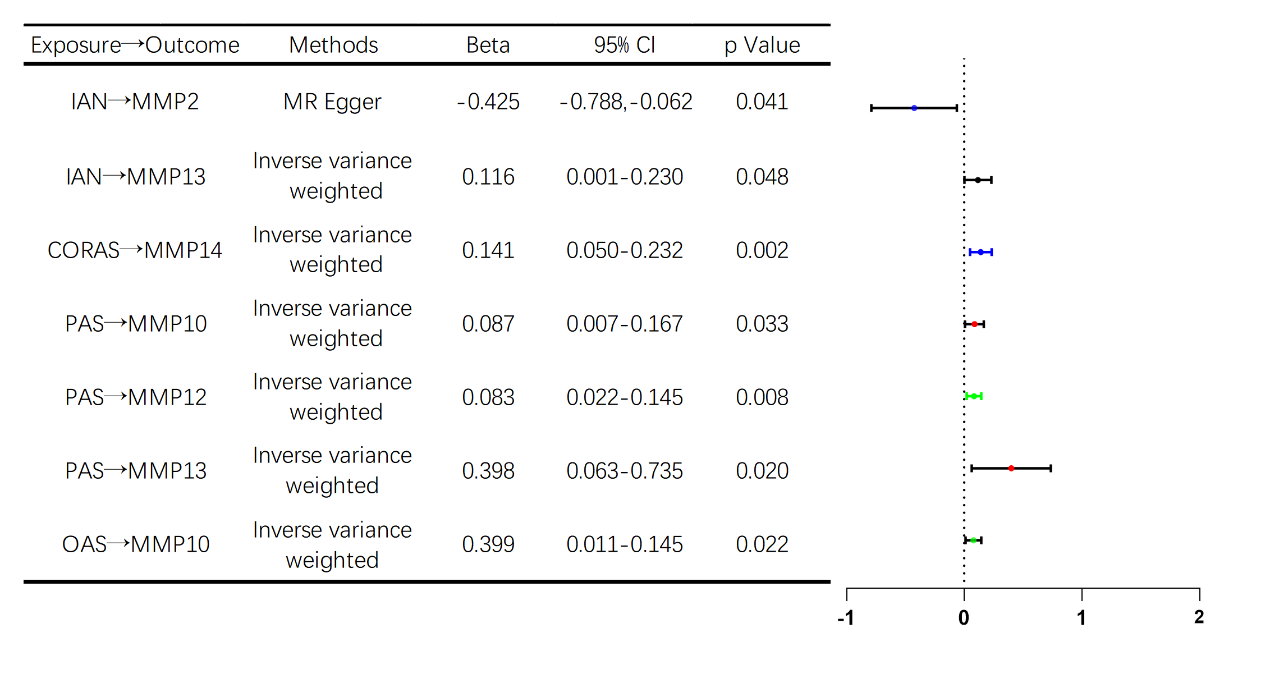

**Supplementary Figure 4.** Forest plot showing changes in circulating MMP family levels due to intracranial aneurysms and atherosclerosis (P < 0.05).

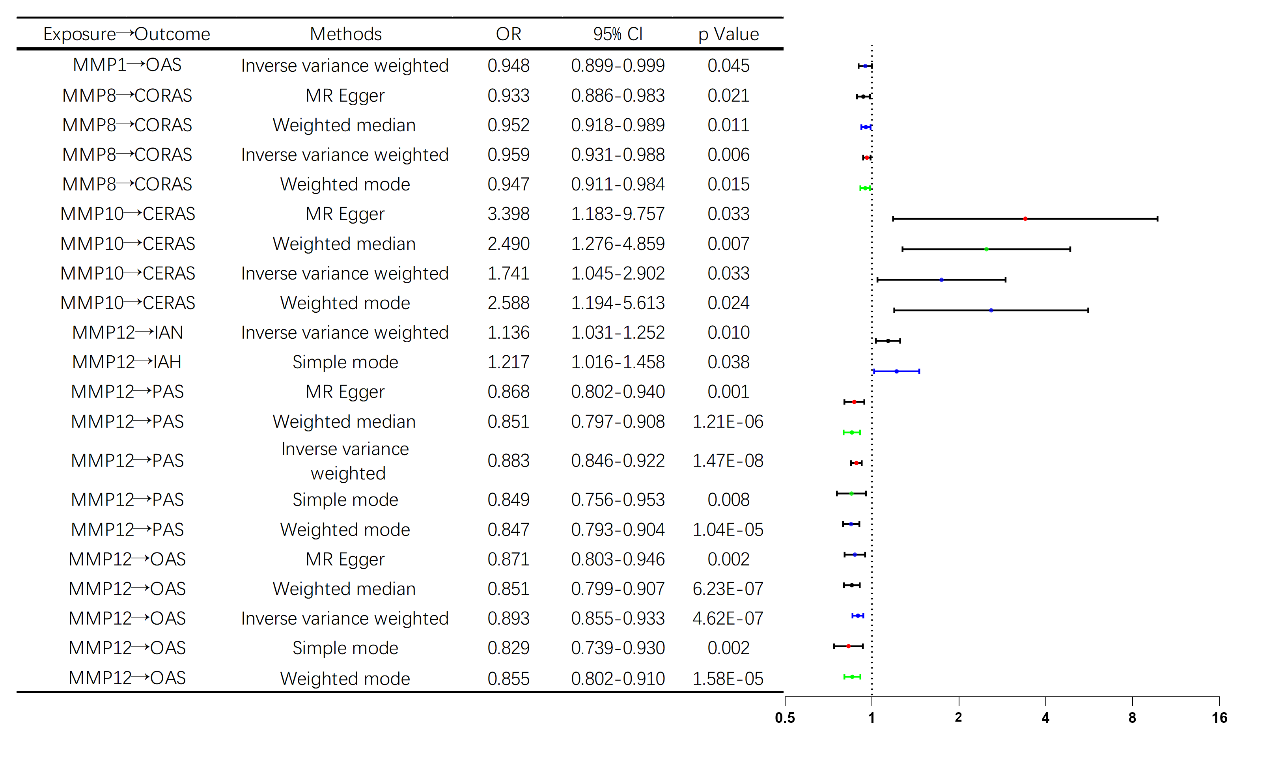

**Supplementary Figure 5.** Forest plot showing changes in the risk of intracranial aneurysms and atherosclerosis due to circulating MMP family levels (P < 0.05).
