## supplementary materials for "Atherosclerosis, Intracranial Aneurysms, and Intermediate Biomarkers: Real-World Observational and Mendelian Randomization Research"

**1.Data Sources of Mendelian Randomization**

**1.1 Genetic instrumental variables for intracranial aneurysm and atherosclerosis**

We extracted genetic association data for intracranial aneurysms and atherosclerosis from genome-wide association studies (GWAS) conducted on three Finnish cohorts. Cases of intracranial aneurysms and atherosclerosis were identified based on inpatient registry, outpatient registry or cause of death.

**1.2 Genetic instrumental variables for matrix metalloproteinases**

We identified the genetic predictors of the 12 matrix metalloproteinases from a comprehensive meta-analysis of plasma protein-associated GWASs from three independent cohorts: Suhre K et al., Folkersen L et al., and Sun BB et al^1-3^. The sample sizes for the GWASs of all MMPs are shown in Table 2. Notably, there was no sample overlap between the MMPs and intracranial aneurysm or atherosclerosis datasets, as they were obtained from different consortia.

**2. Validation of plasma ELISA**

To validate the matrix metalloproteinases identified through Mendelian randomization analysis, we studied 38 patients with carotid artery stenosis and 44 patients with intracranial aneurysms who underwent surgical procedures at the Second Neurosurgery Ward of Beijing Tiantan Hospital between July 2022 and November 2023. Blood samples (5 ml) were collected from all participants via venipuncture before surgery. Additionally, 55 healthy individuals, matched for age and gender, were recruited, and blood samples were collected from them as well. The collected blood samples were centrifuged at 4000 rpm for 10 minutes, and the resulting supernatant was stored at -80°C. Subsequent analysis was performed using the MMP12 ELISA detection kit (Abcam, Cat#ab213811). The procedure involved adding 100 µL of standard or plasma to the ELISA plate, followed by incubation at 37°C for 90 minutes. Subsequently, 100 µL of biotinylated antibody was added to all wells and incubated at 37°C for 60 minutes. After three washes with PBS, 100 µL of ABC working solution was added and incubated for another 30 minutes. Following a final wash, 100 µL of substrate solution was added and incubated for 20 minutes in the dark at room temperature. The reaction was stopped by adding 50 μL of stop solution, and the protease concentration was quantified by recording the absorbance at 450 nm.

**3. Intracranial Aneurysm Mouse Model**

The left kidney of 10-week-old C57/6j mice was surgically removed, and the left common carotid artery was ligated. One week later, 2.5 µL of 35 mU of elastase (Sigma, E1250) was precisely injected into the right basal cistern using a Hamilton microsyringe (26G). The injection site was located 1.2 mm rostral and 0.7 mm lateral to the bregma. Doca sustained-release tablets (Innovative Research of America; M-121, 50 mg) were surgically implanted in the neck, and the mice were provided with 1% sodium chloride drinking water postsurgery. Following the induction of intracranial aneurysms, the mice were closely monitored for signs of hemiplegia, abnormal limb posture, or significant weight loss (>2 g in a day), which could indicate aneurysm rupture. If such signs were observed, the mice were immediately perfused with gelatin and 1% bromophenol blue solution through the left ventricle. Mice without neurological symptoms were euthanized using the same method on the 21st day postinduction. An aneurysm was defined as a localized bulge or dilation in any cerebral artery that was at least 1.5 times the diameter of the parent artery^4,5^.

**5. Local tissue immunostaining**

The stenotic intimal plaque of the carotid artery removed during carotid endarterectomy was fixed in formalin solution for 3-5 days. Subsequently, the plaque tissue was decalcified in a solution (ZLI-9307, Zhongshan Jinqiao, Beijing) for more than 72 hours. The tissue was then gradually immersed in a series of ethanol solutions, starting from 75% ethanol and progressing to absolute ethanol. Each step involved soaking at room temperature for more than 12 hours. Following immersion in xylene, the tissue was embedded in liquid paraffin at 60°C for 2-3 hours and then sectioned to a thickness of 6 µm. The sections were categorized based on the distal normal intima and proximal plaque. Further processing involved deparaffinization, antigen retrieval in boiling ethylenediaminetetraacetic acid (EDTA, pH 8.0) for 15 minutes, and subsequent washing in xylene, graded ethanol, and buffered saline. Specific polyclonal rabbit-derived antibodies against MMP12 (Abcam, Cat# ab128030) were used at a 1:100 dilution for staining. The sections were incubated with primary antibody overnight at 4°C, followed by the addition of secondary antibodies in PBS at a 1:400 dilution at room temperature for 60 minutes. 3,3′-Diaminobenzidine (DAB, Zhongshan Jinqiao) was used as the chromogenic reagent. The reaction was terminated with double-distilled water after a 10-minute incubation at room temperature. Nuclei were stained with hematoxylin, dehydrated step by step, and clarified with xylene. The stained pathological sections were then sealed with gum and photographed using a multispectral scanner.

Aneurysm model mice and age-matched control mice were sacrificed. The brains were removed, fixed in formalin solution, embedded in paraffin, and sectioned to a thickness of 6 µm. After dewaxing and antigen retrieval, the tissue sections were stained using specific rabbit polyclonal antibodies against MMP12 (Thermo Fisher, Cat# PA5-13181) and rat polyclonal antibodies against CD31 (Thermo Fisher, Cat#11-0311-82), both of which were diluted 1:200. The sections were incubated with primary antibody overnight at 4°C, followed by the addition of secondary antibodies in PBS at a 1:600 dilution at room temperature for 60 minutes. The slides were then mounted with DAPI-containing glycerol for nuclear staining and photographed under a multispectral scanner.
